# Predictors of Survival among 6-59 Months Old Children with Severe Acute Malnutrition: A Retrospective Cohort

**DOI:** 10.1101/2022.04.25.22274288

**Authors:** Mohammed Abate, Abel Gebre, Bisrat Tamene Bekele, Fantaye Teka Dinkashe

**Affiliations:** Health System Strengthening and Special Support Directorate, Ministry of Health Ethiopia; Department of Public Health, College of Medical and Health Sciences, Samara University; Monitoring, Evaluation and Quality Improvement, Zewditu Memorial Hospital; St. Paul’s Hospital Millennium Medical College

**Keywords:** Severe Acute Malnutrition, Survival, Hazard Ratio

## Abstract

**Background:** Undernutrition among children is a significant contributor to the global disease burden and a leading cause of child mortality. Ethiopia, home to more than 16 million children under 5 years old, is one of the countries that have high levels of wasting. The aim of this study was to assess survival status and predictors of mortality among children with severe acute malnutrition admitted to Dubti Zonal Referral hospital from January 1/2017 to September 30/2019.

**Methods:** Facility-based retrospective cohort was conducted among 331 severely acutely malnourished children. Data were collected from SAM management registration, individual patient cards and multi-charts admitted from January 1/2017 to September 30/2019. Cox-regression was used to characterize survival within the cohort and to estimate the effect of specific variables while controlling for potential confounders. The hazard ratio was used as a measure of the outcome. P-value less than 0.05 was considered statistically significant to identify independent predictors in multivariable analysis.

**Result:** The median age of study participants was 18 months and males were 187(56%). About half of 160 (48.3%) respondents were with co-morbidities during admission: diarrhea (44%) and pneumonia (26%) were the major co-morbidities. From a total of 331 SAM children, 255(77%) were recovered, 34(10%) died, and 40(12%) have defaulted from treatment. The main risk factors for earlier death of severely malnourished children were rural residence (AHR=1.6, 95% CI= 0.745-3.493), being on IV Infusion (AHR=2.5, 95% CI= 1.12-4.18), anemia during admission (AHR= 6.27, 95% CI =2.41-16.36) & pneumonia (AHR=0.27, 95% CI = 0.11-0.68)

**Conclusions:** The death rate was 10% which is close to the minimum SPHERE standard & national management protocol for SAM. Predictors for earlier hospital deaths were rural residence, IV infusion, Anemia, and Pneumonia.

## Background

Severe acute malnutrition is defined as the presence of oedema of both feet or severe wasting (weight-for-height/length <-3SD or mid-upper arm circumference < 115 mm) (1). Children’s age under 59 months is the critical period for rapid physical growth as well as overall child development and prone to malnutrition. Undernutrition among children is a significant contributor to the global disease burden and a leading cause of child mortality worldwide (2).

The consequences of malnutrition are serious and life-long, falling hardest on the very poor and on children (3). Ethiopia, home to more than 16 million children under 5 years old, is one of the countries that have high levels of wasting (4). In Ethiopia, the prevalence of wasting decreased from 2005 to 2019, from 12% to 7% though it’s higher than the regional average (5,6). The percentage of underweight children has consistently decreased though 7% of children are still wasted, and 1% are severely wasted according to the latest demographic and health survey (6,7).

Regional variations exist, where the Afar region is the second-highest burden (14%) next to Somali (21%) (6,7). A small area estimation of child undernutrition in Ethiopian woredas conducted in 2017 indicated most of Afar region woredas fall under a range of 0.41-1 share of underweight children (8).

Ethiopia launched the first-ever Food and Nutrition Policy in 2018 and the revised guideline for the management of acute malnutrition in 2019 which is expected to provide SAM children an improved quality of care (9,10). Recent studies conducted in Ethiopia indicated a 75.4% to 79% recovery rate (11,12).

Studies in Ethiopia identified that breastfeeding and comorbidities at admission (11) TB, IV fluid infusion, and pale conjunctiva (12) age and vaccination (13) are an independent predictors of death in severely acutely malnourished children. There is no study conducted in Afar region on the magnitude and predictor of death among children admitted for severe acute malnutrition management. This study aimed to assess survival status and predictors of mortality among children with severe acute malnutrition admitted to Dubti Zonal Referral hospital (DZRH) from January 2017 to September 2018.

## Methods

### Study Area and Period

The study was conducted in Dubti zonal referral Hospital, Awssiresu zone, Afar regional state, located 570 km from Addis Ababa. Awssiresu zone has an estimated population of about 613,944 among this about 69,990 are under-five children. There were a referral hospital, a district hospital, twenty health centers and 109 health posts. The community is a pastoralist and prone to recurrent food insecurity. Data were collected from November 2018 to February 2^nd^, 2019.

### Study design

Facility-based retrospective cohort study was conducted. All 6-59 months old children with severe acute malnutrition admitted to Dubti zonal referral Hospital from January 1/2017 to September 30/2019 were included in the study.

### Sample Size Determination and Sampling Procedure

The sample size was calculated by Epi info version 7 statistical software, considering: 95% CI, 80% power, ratio of unexposed to exposed 1:1, outcome (death in this case) for unexposed group 3.46% and for exposed group 11.85% (14). Three hundred ninety-six samples were selected by using simple random sampling techniques from eligible participants listed on the admission ward registration book through a computer program.

### Data collection tools and procedures

The data collection checklist was developed from the current standard treatment protocol for the management of severe acute malnutrition (10). Data were collected using the pretested (at Asayita hospital) checklist by three diploma clinical nurses from SAM management registration, individual patient cards and multi-charts supervised by a BSc nurse/public health supervisor.

### Data quality control

Training was provided for data collectors and supervisors on the objective and data collection tools with a focus on how and what information to be collected. The completeness and consistency of the collected data were checked daily by the supervisors and the principal investigator. Whenever there appears incompleteness and ambiguity of recording, the filled information formats were crosschecked with source data. Individual records with incomplete data were excluded.

### Operational definitions

**Co-morbidities**: children with severe acute malnutrition, who had diarrhea, TB, and/or HIV and/or malaria and/or severe anemia co-infection at admission to DZRH.

**Dead**: Patient that has died, while he/ she was in the program recorded by health worker.

**Defaulter**: Patient that is absent for 2 days in in-patient.

**Recovered;** Patient that has reached the discharge criteria during treatment at stabilization center.

**Severe malnutrition**: weight-for-height z scores (WHZs) of less than or equal to −3 or less than or equal to 70% of the reference median of WHO reference values (severe wasting) or symmetrical edema involving at least the feet(1).

**Exposed children**: All severely malnourished children aged 6-59 months with Co-morbidities admitted to TFU in hospital.

### Data analysis and processing

Data were cleaned and coded before data entry to Epi data version 3 and exported to STATA for analysis. Patients admitted to stabilization centers in the hospital who are recovered, defaulted, transfer, medical referral, and unknown were considered as censored observation whereas death is an event of interest in this study. Time until death was computed using STATA software by subtracting admission date from discharge date.

Survival status (dependent variable) recode into dichotomies variables; alive (censored observation) and died (event), and valued (1 if censored, 2 if died) before conducting analysis. Univariable analysis was conducted and presented using tables and figures. Bivariable analysis was done using Cox-regression to identify crude associations between dependent and independent variables and to select variables for multivariable analysis.

Before conducting multivariable Cox-regression, multicollinearity among variables was checked. Cox regression model assumption of proportional hazards was checked by testing the interaction of covariates with time. Kaplan Meier was used to estimate mean survival time during the treatment period. Variables significant at P <0.25 level in the Bivariable analysis were included in the final Cox-regression analysis. Multivariable Cox regression was run using Forward Wald (stepwise) method to identify the best independent predictors of earlier death. Statistical significance was set at a P value less than 0.05.

## Results

### Socio-demographic characteristics

Out of 396 randomly selected SAM children records the data of 331(83%) were eligible to be included and all necessary data were extracted. The median age of study participants was 18 months (range 6-24). Males were 187(56%) and most 182(55%) of the study participants live in rural areas (Table **1**).

**Table 1:** Socio demographic and Clinical characteristics of severely malnourished 6-59 months of children admitted to DZRH, Afar regional state, 2019.

| Variable | Categories | Outcomes |  |  |
| --- | --- | --- | --- | --- |
|  |  | Censored (n, %) | Death (n, %) | Total (n, %) |
| Sex | Male | 158(84.5%) | 29(15.5%) | 187(56%) |
|  | Female | 139(96.5%) | 5(3.5%) | 144(44%) |
| Age | 6 -24 month | 209(91%) | 21(9%) | 230(69%) |
|  | ≥24 month | 88(87%) | 13(13%) | 101(31%) |
| Residence | Rural | 157(86.3%) | 25(13.7%) | 182(55%) |
|  | Urban | 140(94%) | 9(6 %) | 149(45%) |
| Vomiting | Present | 86(29%) | 15(44%) | 101(31%) |
|  | Absent | 211(71%) | 19(56%) | 230(69%) |
| Temperature | Altered | 70 (24%) | 14(41%) | 84(25%) |
|  | Normal | 227(76%) | 20(59%) | 247(75%) |
| Respiration | Altered | 67(23%) | 16(47%) | 83(25%) |
|  | Normal | 230(77%) | 18(53%) | 248(75%) |
| Pulse Rate | Altered | 21(7%) | 5(15%) | 26(8%) |
|  | Normal | 276(93%) | 29(85%) | 305(92%) |
| Edema | Present | 33(11%) | 5(15%) | 38(11%) |
|  | Absent | 264(89%) | 29(85%) | 293(89%) |
| Palmer pallor | Present | 35(12%) | 4(12%) | 39(12%) |
|  | Absent | 262(88%) | 30(88%) | 292(88%) |
| Skin lesion | Present | 2(0.7%) | 2(6%) | 4(1.2%) |
|  | Absent | 295(99.3%) | 32(94%) | 227(98.8) |
| Dehydration | Present | 50(16%) | 12(44%) | 62(19%) |
|  | Absent | 247(84%) | 22(56%) | 269(81%) |
| Shock | Present | 2(0.7%) | 2(6%) | 4(1.2%) |
|  | Absent | 295(99.3%) | 32(94%) | 227(98%) |
| Co-morbidity | Present | 127(43%) | 33(97%) | 160(48%) |
|  | Absent | 170(57%) | 1(3%) | 171(52%) |
| Conjunctiva color | pink | 35(12%) | 4(12%) | 39(12%) |
|  | pale | 262(88%) | 30(88%) | 292(82%) |
| Consciousness level | conscious | 293(99%) | 31(91%) | 324(98%) |
|  | Impaired | 4(1%) | 3(9%) | 7(2%) |
| MUAC | <11.5cm | 207(70%) | 22(65%) | 229(69%) |
|  | ≥11.5cm | 90(39%) | 12(35%) | 102(31%) |

### Co-morbidity and Clinical Conditions

About half of 160 (48.3%) respondents presented with co-morbidities during admission; among them, diarrhea (44%) and pneumonia (26%) were the major co-morbidities (Figure 1). Among the children with diarrhea, 62(19%) of them were dehydrated and MUAC measurement <11 was 22(66.6%) (Table **1**).

**Figure 1:**
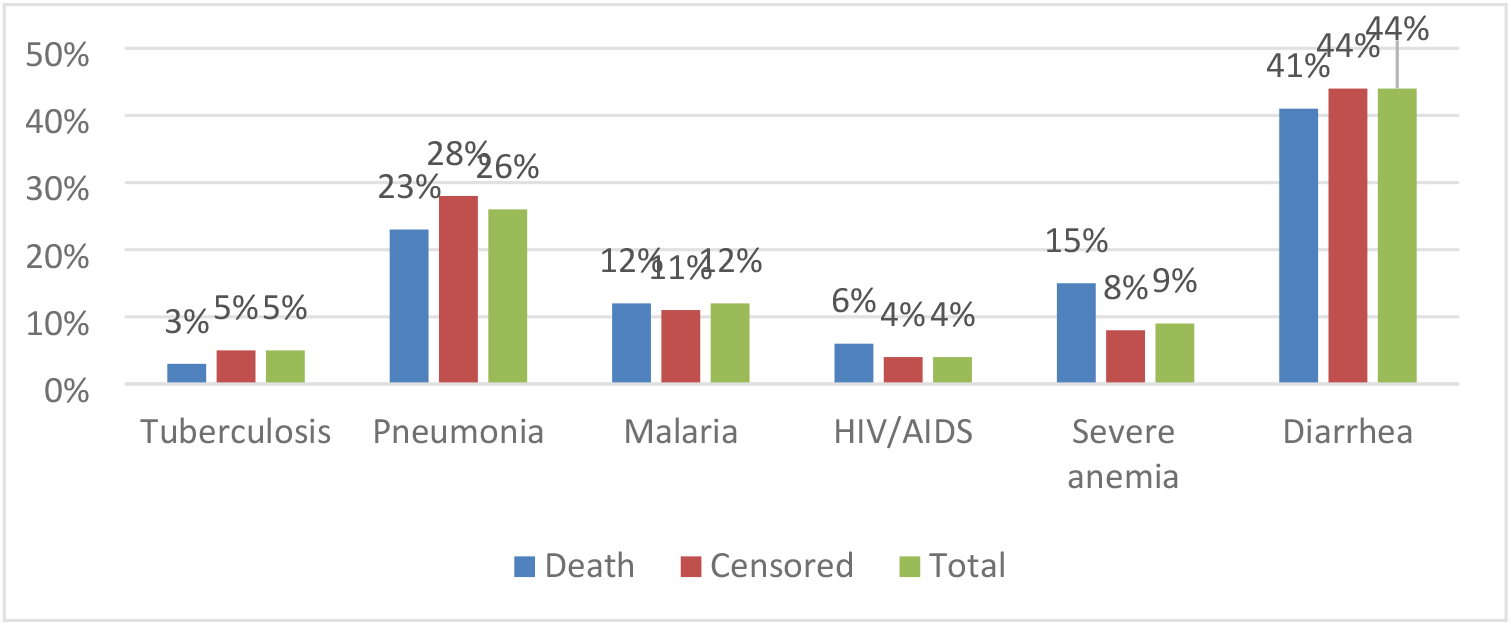
Co-morbidities at admission among severely malnourished children admitted to DZRH, Northeast Ethiopia, 2019.

### Types of treatment given

From the total children who got treatment for SAM in DZRH, 97% were on Intravenous (IV) antibiotics while 82% were supplemented with vitamin A. Only one percent of the study participants were infused which was computed as six percent of the dead during treatment (Figure 2).

**Figure 2:**
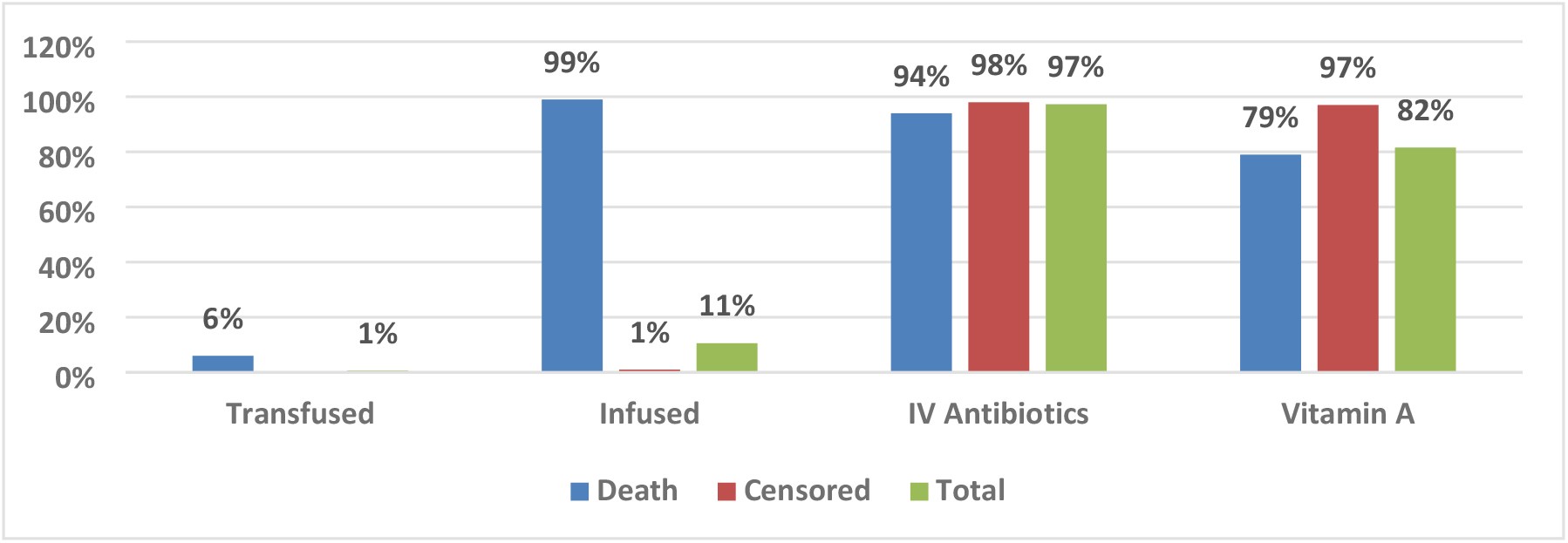
Type of treatment given of severely children admitted to DZRH, Northeast Ethiopia, 2019.

### Treatment Outcomes

All study participants were followed for a median of 10 days (range 1-22) which gives 2334-person days of observation; the pick discharge with the improved and required nutritional transfer. A total of 255 (77%) were improved and required nutritional transfer, and 34(10%) died during treatment. Among those transferred, about three-fourth (77%) of them transferred from the stabilization center at the end of the first week. Sixty-six percent were discharged in the first week while 29% were discharged in the second week and 5% in the third week.

Out of 255 children discharged with improvement, only 79 (31%) of the children achieved a target weight of 85% weight for height. From all deaths occurred during the treatment period in the stabilization center, 18% occurred within two days, 38% occurred within 7 days and 44% occurred in the second week from date of admission. The assessment showed overall mean survival time was 18.39 (95% CI=17.196-19.514). However, the mean survival time for children presented with co-morbidity at admission was lower than compared to children without comorbidities, 15.41(13.57-17.26) and 20.55(19.69-21.41) respectively (Figure 3). According to the hazard function, children with comorbidities were more likely to die at any time during follow up than children without comorbidities (Figure 4).

**Figure 3:**
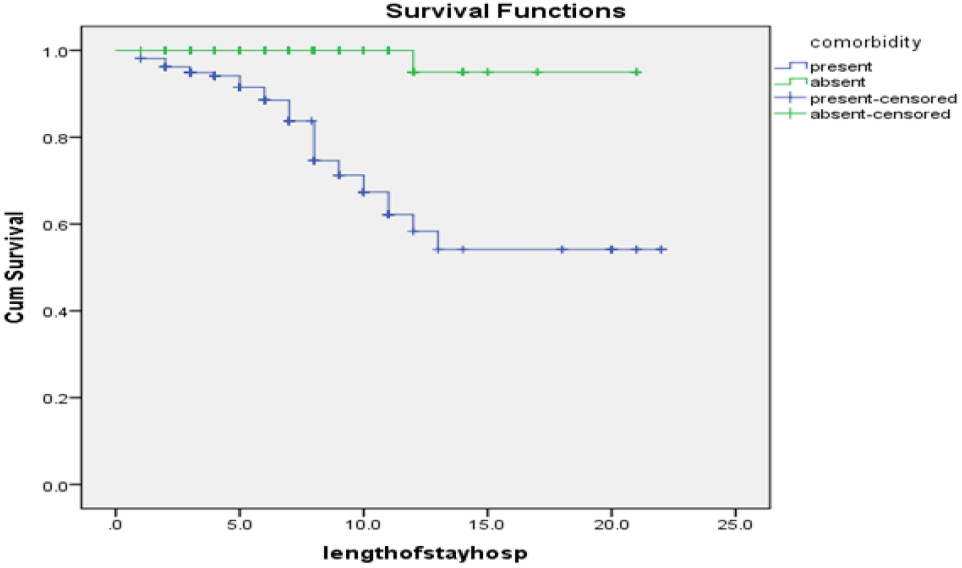
The Kaplan-Meier survival function difference of SAM children admitted with and without co-morbidity admitted to DZRH, Northeast Ethiopia, 2019.

**Figure 4:**
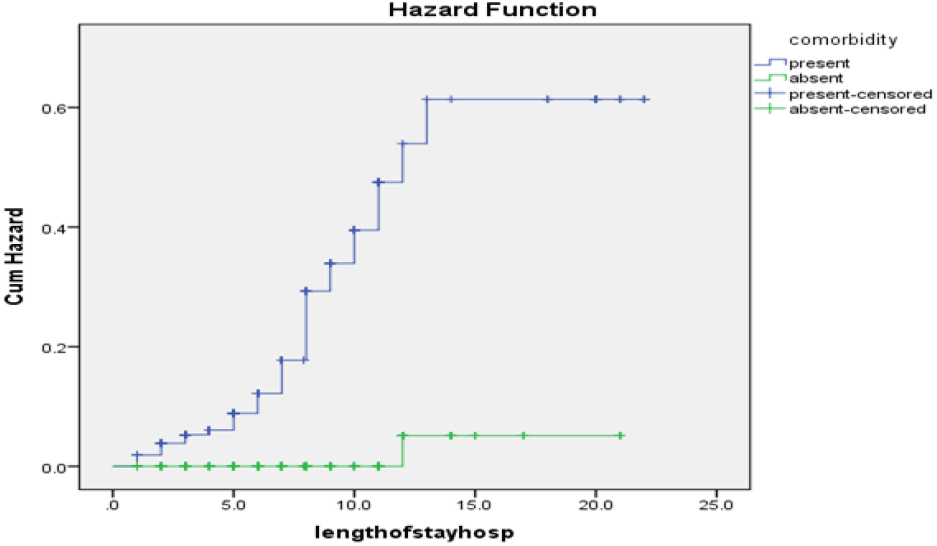
The Kaplan Meier hazard function difference of death among severely malnourished children admitted to DZRH, Northeast Ethiopia, 2019.

### Predictors of Mortality among SAM children

Bivariable analysis was done for all independent variables using Cox-regression for socio-demographic characteristics (sex, age and resident), underlying clinical conditions, co-morbidity at admission, and type of treatment given. Residence, vomiting, Anemia, pneumonia, shock, fluid infusion and consciousness were found to be candidates for multivariable analysis at a P value less than 0.25.

Multivariable Cox regression was run for variables found to be candidates in Bivariable Cox regression. Before regression, overall model fitness was checked by likelihood and chi-square test. Regression was run using the Backward Wald method to identify the best independent predictors of earlier death. A P-value of less than 0.05 was used as statistical significance. After performing multivariable Cox-regression analysis, variables that have a significant level at 95% CI and P value < 0.05 were considered in predicting earlier mortality. Residence, fluid infusion, pneumonia, and Anemia, were found to be independent predictors of death in severely malnourished under-five children admitted to DZRH (The **risk** of mortality among children with SAM admitted to SCs from the rural areas was 1.61 times higher as compared to urban residents (AHR= 1.61, 95% CI 0.74–3.49). The hazard of mortality among children with SAM admitted to SCs having anemia was 6.28 times higher as compared to non-anemic children (AHR = 6.28, 95% CI 2.41-16.36). Children who were on IV infusion during admission were 2.51 times at risk of mortality as compared to children who did not (AHR=2.51, 95% CI 1.12-4.18 (Table **2**).

Table 2)

**Table 2:** Predictors with survival status of severely malnourished children of under-five years admitted to DZRH, Northeast Ethiopia, 2019.

| Predictor variable | Categories | Treatment Out come |  |  |  |  |  |
| --- | --- | --- | --- | --- | --- | --- | --- |
|  |  | Event/ Death | Censored | CHR<br>95 % CI | P value | AHR (95%CI) | P value |
| **Resident** | Rural | 25 | 157 | 2.61(1.12-6.06) | 0.026 | 1.61(.74-3.49) | 0.002 |
|  | Urban | 9 | 140 | 1 |  |  |  |
| **IV Infused** | Yes | 32 | 3 | 5.7 (4.85-28.33) | 0.001 | 2.51(1.12-4.18) | 0.001 |
|  | No | 2 | 294 | 1 |  |  |  |
| **Anemia** | Yes | 5 | 10 | 7.44(2.39-23.16) | 0.010 | 6.28(2.41-16.36) | 0.001 |
|  | No | 29 | 287 | 1 |  |  |  |
| **Pneumonia** | Yes | 8 | 35 | 2.54(1.15-5.64) | 0.021 | 0.27(0.11-0.68) | 0.005 |
|  | No | 26 | 262 | 1 |  |  |  |

The risk of mortality among children with SAM admitted to SCs from the rural areas was 1.61 times higher as compared to urban residents (AHR= 1.61, 95% CI 0.74–3.49). The hazard of mortality among children with SAM admitted to SCs having anemia was 6.28 times higher as compared to non-anemic children (AHR = 6.28, 95% CI 2.41-16.36). Children who were on IV infusion during admission were 2.51 times at risk of mortality as compared to children who did not (AHR=2.51, 95% CI 1.12-4.18 (Table **2**).

## Discussions

In this study, SAM children were followed for a median of 10 days (range 1-22). A closer average length of stay in the stabilization center was reported in the Tigray region (northern Ethiopia), 12 days, and in the Gedeo zone (southern Ethiopia), 14 days (15,16). However, this is much lower than the international standard (SPHERE) set for the management of SAM which recommends no more than 30 days (17). The difference can be explained by the clinical profiles of children, chronic commodities like TB and HIV/ADIS might need more time in hospitals (10).

Out of 255 children discharged with improvement, only 79(31 %) of the children achieved a target weight of 85% weight for height. Similarly, in Jimma university’s specialized hospital, only 226 (30.6 %) of the children had achieved the target weights (18).

In this study Diarrhea (42%) was the most prevalent co-morbidity that occurred among severely malnourished children. A similar finding (44.6%) was reported from Sekota hospital, Northwest Ethiopia (14). Diarrhea incidence is still high though the government endeavor to increased access to a safe source of drinking water and improved sanitation facilities according to the 2016 EDHS (7).

The finding of this study revealed that 10% of the children died during the follow-up period which is at the borderline with the minimum SPHERE standard and national management protocol for severe acute malnutrition managed at stabilization centers (< 10%) (17). This finding is similar to other assessments conducted in Ethiopia: Felege-hiwot comprehensive specialized hospital, Gedeo Zone and Jimma University specialized hospital (19) (20) (18). On the contrary, a low death rate was reported in the south Wollo zone (3.4%), the Amhara region and the Tigray region (3.8%) in Ethiopia (11,15). On the other side, the result is lower than the study conducted in Sekota hospital (28.7%) (14).

In this assessment, from all deaths that occurred during the treatment period, 18% of them occurred within 48 hours, 38% of them occurred within 7 days. A higher percentage was reported from Jimma for both weeks; Of the 88 deaths, 27.3 % occurred in the first 48 h and 60.2 % by the end of the first week (18). This variation might be due to differences in space availabilities, patient load, patient profile, medical team and medical supplies.

In this study, residence, pneumonia, and anemia were found to be independent predictors of death. Similarly, other investigations in Zambia and South Africa (21,22) and in Ethiopia 14, 16,18,23) showed that there was a significant association between SAM child mortality and pneumonia or anemia. Another study in Tigray reported being urban in residence was one of the independent predictors of mortality (15). However, a study conducted in Felegehiwot hospital did not show any significant association between death of severe acute malnutrition children with pneumonia, shock, TB, type of SAM, diarrhea, anemia, CHF (19).

This discrepancy probably be because of difference in sample size or might be the difference in the study period as there were changes in treatment modality and updating of professionals in standard training and regular supervision. The other possible justification could be strictly using the national management protocol, for example, TB and SAM management guidelines (19).

The risk of mortality among children with SAM admitted to stabilization center from the rural areas were 1.61 times at higher risk of death compared to urban residents. In Tigray region, reverse result was reported where SAM children from urban areas were 2.73 times at higher risk of death compared to rural residents (15). This can be explained by the majority (74.8%) of the children in the study were from the rural areas and the total number of deaths observed is small as compared to the number of children.

The hazard rate of mortality among children with SAM presented with anemia was 6.28 times at higher risk of death compared to non-anemic children. Similar study conducted in Gedeo Zone, reported 2.62 times higher risk of death rate among children with anemia while 2.1 times was from Jimma University specialized hospital (18, 20). A research conducted in Sekota hospital also showed the case fatality rate of children with severe anemia was higher (14). This could be explained by during anemia the prevalence of infection and the probability of heart failure increases while compliance will decrease.

Children who had IV infusion during admission were 2.51 times at risk of mortality as compared to children who did not. A study in Addis Ababa and Dilla reported IV fluid infusion was independent predictor of mortality (12,16). In another study conducted in Jimma, treatment related factors like infusion and transfusion were not independent predictors of death in severely malnourished children (18).

Other disease such as TB & malaria did not predict mortality in our study. However, in another study the risk of death due to TB was three fold & that ascribed to malaria was 2.13 fold (95% CI=1.12-7.35) this may be due to that the hospital was properly managed & majority of malaria cases were mild (19).

## Conclusion

Based on the finding of study, only 31% gained wt/ht >85%, during discharge. Death rate was 10% and risk factors or predictors for earlier hospital deaths among severely malnourished children admitted to a hospital were rural residence, IV infusion, Anemia and Pneumonia.

## Limitation of the study

Since the data were retrospectively collected from SAM Children’s medical records, some important variables were excluded such as, parents’ socio-demographic characteristics. In addition, incomplete records were excluded from analysis.

## Data Availability

The data underlying the results presented in the study are available from Mohammed Abate Health System Strengthening and Special Support Directorate, Ministry of Health Ethiopia Email-

## Abbreviations

SAM: Severe acute malnutrition
Wt: weight
Ht: Height
IV: Intravenous
WHZ: Weight for Height-Z score

## Declarations

### Ethical considerations

Ethical clearance was obtained from Institutional Review Board of Samara University Health Science College. Prior to data collection, permission was secured from Afar Regional Health Bureau and DZRH. The data were kept confidential and used for the intended purpose only.

### Consent for publication

Not applicable

### Availability of data and materials

The datasets used and/or analyzed during the current study are available from the corresponding author on reasonable request.

### Authors’ contributions

MA and AG designed the study and analyzed the data. BTB and FTD wrote the final report and drafted the manuscript. All authors read and approved the final manuscript

## Acknowledgements

The authors would like to acknowledge study participants, data collectors and administrative bodies for their permission.

## References

1. World Health Organization. Pocket Book of Hospital Care for Children: Guidelines for the Management of Common Childhood Illnesses. 2nd ed. Geneva, Switzerland; 2013.

2. Black RE, Allen LH, Bhutta ZA, Caulfield LE, De Onis M, Ezzati M et al. Maternal and child undernutrition: global and regional exposures and health consequences. Lancet. 2008;371(9608):243–60.

3. United Nations Children’s Fund, World Health Organization, International Bank for Reconstruction and Development WB. Levels and trends in child malnutrition: key findings of the 2018 Edition of the Joint Child Malnutrition Estimates. Geneva, Switzerland; 2018.

4. Laillou A, Baye K, Meseret Z, Darsene H, Rashid A, Chitekwe S. Wasted children and wasted time: A challenge to meeting the nutrition sustainable development goals with a high economic impact to ethiopia. Nutrients. 2020;12(12):1–12.

5. UNICEF, WHO WBG. LEVELS AND TRENDS IN CHILD MALNUTRITION, UNICEF / WHO / World Bank Group Joint Child Malnutrition Estimates Key findings of the 2020 edition [Internet].2020. Available from: https://www.who.int/publications/i/item/jme-2020-edition

6. Ethiopian Public Health Institute (EPHI) [Ethiopia] and ICF. Ethiopia Mini Demographic and Health Survey 2019: Key Indicators. Rockville, Maryland, USA:; 2019.

7. CSA, ICF. Ethiopia Demographic and Health Survey 2016. [Internet]. Addis Ababa, Ethiopia, and Rockville, Maryland, USA,; 2016. Available from: https://dhsprogram.com/pubs/pdf/FR328/FR328.pdf

8. Sohnesen TP, Ambel AA, Fisker P, Andrews C, Khan Q. Small area estimation of child undernutrition in Ethiopian woredas. PLoS One. 2017;12(4):1–17.

9. Federal Democratic Republic of Ethiopia. Ethiopia Food and Nutrition Policy. Food and Nutrition Policy. Addis Ababa, Ethiopia; 2018.

10. Federal Ministry of Health. National Guideline for the Management of Acute Malnutrition in Ethiopia. National Guideline for the Management of Acute Malnutrition in Ethiopia. Addis Ababa, Ethiopia; 2019.

11. Hassen SL, Astatkie A, Mekonnen TC, Bogale GG. Survival Status and Its Determinants among Under-Five Children with Severe Acute Malnutrition Admitted to Inpatient Therapeutic Feeding Centers in South Wollo Zone, Amhara Region, Ethiopia. J Nutr Metab. 2019;2019.

12. Bitew ZW, Alebel A, Worku T, Alemu A. Recovery rate and its predictors among children with severe acute malnutrition in Addis Ababa, Ethiopia: A retrospective cohort study. PLoS One [Internet]. 2020;15(7 July). Available from: http://dx.doi.org/10.1371/journal.pone.0235259

13. Desyibelew HD, Baraki AG, Dadi AF. Mortality rate and predictors of time to death in children with severe acute malnutrition treated in Felege-Hiwot Referral Hospital Bahir Dar, Northwest Ethiopia. BMC Res Notes [Internet]. 2019;12(1). Available from: https://doi.org/10.1186/s13104-019-4467-x

14. Shitaye Desta K. Survival Status and Predictors of Mortality among Children Aged 0-59 Months with Severe Acute Malnutrition Admitted to Stabilization Center at Sekota Hospital Waghemra Zone. J Nutr Disord Ther. 2015;5(2):1–11.

15. Guesh G, Degu G, Abay M, Beyene B, Brhane E, Brhane K. Survival status and predictors of mortality among children with severe acute malnutrition admitted to general hospitals of Tigray, North Ethiopia: A retrospective cohort study. BMC Res Notes [Internet]. 2018;11(1):1–7. Available from: https://doi.org/10.1186/s13104-018-3937-x

16. Adal TG, Kote M, Tariku B. Incidence and predictors of mortality among severe acute malnourished under five children admitted to Dilla University Referal hospital: a retrospective longitudinal study. J Biol Agric Healthc. 2016;13(May).

17. The Sphere Project. Humanitarian Charter and Minimum Standards in Disaster Response [Internet]. Geneva, Switzerland; 2004. Available from: https://www.refworld.org/pdfid/3d64ad7b1.pdf

18. Jarso H, Workicho A, Alemseged F. Survival status and predictors of mortality in severely malnourished children admitted to Jimma University Specialized Hospital from 2010 to 2012, Jimma, Ethiopia: A retrospective longitudinal study. BMC Pediatr [Internet]. 2015;15(1):1–13. Available from: http://dx.doi.org/10.1186/s12887-015-0398-4

19. Kassaw A, Amare D, Birhanu M, Tesfaw A, Zeleke S, Arage G, et al. Survival and predictors of mortality among severe acute malnourished under-five children admitted at Felege-Hiwot comprehensive specialized hospital, northwest, Ethiopia: a retrospective cohort study. BMC Pediatr. 2021;21(1):1–10.

20. Girum T, Kote M, Tariku B, Bekele H. Survival status and predictors of mortality among severely acute malnourished children <5 years of age admitted to stabilization centers in Gedeo Zone: A retrospective cohort study. Ther Clin Risk Manag. 2017;13:101–10.

21. Munthali T, Jacobs C, Sitali L, Dambe R, Michelo C. Mortality and morbidity patterns in under-five children with severe acute malnutrition (SAM) in Zambia: A five-year retrospective review of hospital-based records (2009-2013). Arch Public Heal. 2015;73(1):1–9.

22. Muzigaba M, Puoane T, … BS-… AJ of C, 2017 undefined. Independent and interactive effects of HIV infection, clinical stage and other comorbidities on survival of children treated for severe malnutrition in rural South Africa: A. AjolInfo [Internet]. 2017;11(1):46–53. Available from: https://www.ajol.info/index.php/sajchh/article/view/154499

23. Oumer, A., Mesfin, F., Demena M 2016. Survival Status and Predictors of Mortality among Children Aged 0-59 Months Admitted with Severe Acute Malnutrition in Dilchora Referral Hospital, Eastern Ethiopia. East African Journal of Health and Biomedical Sciences, Volume 1 (1): 13–22. 2016;1(January):13–22.

